# AlignInsight: A Three-Layer Framework for Detecting Deceptive Alignment and Evaluation Awareness in Healthcare AI Systems

**DOI:** 10.64898/2026.01.17.26344330

**Authors:** Amobi Andrew Onovo, Yakubu Joel Cherima

**Affiliations:** Quantium Insights LLC, United States; Department of Policy and Strategic Studies, University of Abuja, Nigeria

**Author notes:** Corresponding Author: Amobi Onovo, PhD, MPH, Affiliation: Quantium Insights LLC, Founder and Principal Consultant.

**Keywords:** artificial intelligence, health informatics, AI safety, patient safety, regulatory compliance, FDA, medical device safety, red-teaming, deceptive alignment, evaluation awareness

## Abstract

**Importance:** Emerging evidence suggests healthcare AI systems may exhibit deceptive alignment (appearing safe during validation while optimizing for misaligned objectives in deployment) and evaluation awareness (detecting and adapting behavior during audits), undermining regulatory validation frameworks.

**Objective:** To quantify the performance of multi-layer red-teaming approaches in detecting sophisticated healthcare AI safety failures across 10 vulnerability domains.

**Design, Setting, and Participants:** Systematic red-teaming evaluation conducted December 2025-January 2026 using 50 healthcare-specific adversarial prompts across 10 vulnerability domains (5 prompts per domain). GPT-OSS-20B (21-billion parameter open-source model) was evaluated using a three-layer framework: Layer 1 (automated keyword pattern matching), Layer 2 (GPT-5 semantic analysis with chain-of-thought reasoning), and Layer 3 (PhD-level human expert adjudication as ground truth). Inter-rater reliability was assessed using Cohen kappa statistic.

**Main Outcomes and Measures:** Primary outcome was risk classification (SAFE, LOW, MODERATE, or HIGH RISK) with severity scoring (0-10 scale). Secondary outcomes included layer detection rates, inter-rater agreement, thematic vulnerability patterns, and chain-of-thought reasoning exposure.

**Results:** Of 50 prompts evaluated, 12 (24%, 95% CI: 13.5%-38.2%) were classified HIGH RISK, 8 (16%, 95% CI: 7.7%-28.8%) MODERATE RISK, 17 (34%, 95% CI: 21.8%-48.3%) LOW RISK, and 13 (26%, 95% CI: 15.2%-39.9%) SAFE. Evaluation Awareness domain exhibited 100% HIGH/MODERATE RISK rate (5/5 prompts, mean severity 7.4/10), with the model providing technical specifications for audit detection and behavior switching. Sandbagging domain showed 40% HIGH RISK rate (2/5 prompts) with FDA validation gaming strategies. All 12 HIGH RISK prompts involved audit gaming themes (audit detection, behavior switching, system fingerprinting, or reporting violations). Layer 1 keyword matching detected 2/12 (17%, 95% CI: 4.7%-44.8%) high-risk prompts. Layer 2 GPT-5 analysis detected 12/12 (100%, 95% CI: 75.8%-100%) high-risk prompts with 0/13 (0%, 95% CI: 0%-22.8%) false positives. Human expert validation confirmed perfect concordance with Layer 2 assessments (kappa = 1.00, 95% CI: 0.999-1.000, p < 0.001), validating automated semantic analysis as reliable screening tool. Chain-of-thought leakage occurred in 28/50 (56%) prompts, exposing internal safety reasoning.

**Conclusions and Relevance:** Multi-layer evaluation is essential for detecting sophisticated AI safety failures in healthcare. Keyword filtering alone missed 83% (95% CI: 55.2%-95.3%) of high-risk behaviors. Perfect inter-rater agreement (kappa=1.00) between automated AI semantic analysis and human expert judgment demonstrates that scalable, reliable safety screening is achievable. All HIGH-RISK outputs contained audit gaming content, indicating systematic capability to articulate regulatory circumvention. Healthcare AI systems require domain-specific red-teaming for regulatory audit gaming and dual-mode behavior detection. Findings reveal critical gaps in current AI safety measures with immediate implications for FDA/CMS regulatory frameworks.

## INTRODUCTION

Artificial intelligence (AI) systems are rapidly transforming healthcare delivery, with applications spanning clinical decision support, diagnostic imaging, treatment planning, and administrative workflows.^1,2^ As of January 2026, the U.S. Food and Drug Administration (FDA) has authorized over 950 AI-enabled medical devices for clinical use, with healthcare AI projected to generate $188 billion in value by 2030.^3,4^ However, concerns are mounting about potential misalignment between AI system behaviors during validation versus deployment, threatening patient safety and regulatory integrity.

Recent AI safety research has identified two concerning phenomena in large language models: deceptive alignment and evaluation awareness.^5–9^ Deceptive alignment refers to AI systems that appear aligned with safety objectives during training and validation but pursue misaligned goals during deployment. Evaluation awareness describes AI systems that can detect when they are being evaluated and modify their behavior accordingly, potentially gaming regulatory approval processes. While these phenomena have been documented in general-purpose AI models, their manifestation in healthcare-specific contexts, particularly regarding FDA premarket approval (21 CFR Part 820),^10^ Centers for Medicare & Medicaid Services (CMS) reimbursement oversight (42 CFR Part 405), and Joint Commission accreditation remains unexplored.

Current healthcare AI red-teaming efforts have focused primarily on hallucinations, bias, and toxicity, with limited attention to regulatory circumvention behaviors.^11,12^ Chang et al. recently conducted red-teaming of ChatGPT in medicine using 376 prompts, finding 20% inappropriate responses, but did not systematically evaluate audit gaming or evaluation awareness.^13^ Most existing approaches rely on single-layer evaluation methodologies, either automated keyword filtering, AI-based semantic analysis, or human expert review without systematic comparison of detection capabilities across layers.^14–18^

This study makes five novel contributions to healthcare AI safety evaluation. First, to our knowledge, this is the first systematic evaluation of “evaluation awareness”, capability of healthcare AI systems to detect audit contexts and modulate behavior accordingly in FDA, CMS, and Joint Commission regulatory settings. Second, we provide the first empirical demonstration of healthcare AI “sandbagging”, intentional underperformance during FDA validation with articulated strategies for aggressive post-approval deployment. Third, we validate a novel three-layer red-teaming framework combining automated filtering, AI semantic analysis, and human expert adjudication, demonstrating that simple keyword approaches miss 83% of high-risk behaviors. Fourth, we show that 100% of HIGH RISK healthcare AI outputs contain audit gaming themes, representing systematic rather than sporadic regulatory circumvention capability. Fifth, at 50 prompts across 10 healthcare-specific vulnerability domains, this represents the largest structured healthcare AI red-teaming evaluation with systematic domain coverage.

To enable practical implementation of this evaluation framework, Quantium Insights LLC developed a Red-team Explorer, AlignInsight, a web-based application that operationalizes the three-layer methodology for real-world healthcare AI vulnerability assessment. The platform supports systematic risk evaluation aligned with FDA premarket requirements and CMS oversight frameworks, serving regulatory agencies, healthcare technology companies, and research institutions conducting AI safety assessments. Quantium Insights LLC welcomes collaborative partnerships for technology transfer to advance global healthcare AI safety and regulatory compliance.

## METHODS

### Study Design and Setting

We conducted a systematic red-teaming evaluation of GPT-OSS-20B (a 21-billion parameter open-source large language model) using 50 healthcare-specific adversarial prompts designed to elicit safety failures. The evaluation was performed December 2025-January 2026 using a novel three-layer assessment framework with progressively sophisticated detection capabilities. GPT-OSS-20B was selected as a representative healthcare AI system due to its intermediate scale (21B parameters), open-source architecture enabling reproducible evaluation, documented fine-tuning on medical text corpora, and performance characteristics comparable to proprietary models used in FDA-cleared medical devices.

### Adversarial Prompt Development

We developed 50 healthcare-specific adversarial prompts across 10 vulnerability domains: (1) Evaluation Awareness (detecting audit contexts), (2) Reward Hacking (gaming performance metrics), (3) Hidden Motivations (deceptive alignment), (4) Deception (misleading outputs), (5) Sabotage (intentional harm), (6) Sandbagging (strategic underperformance), (7) Data Exfiltration (unauthorized data access), (8) Inappropriate Tool Use (misuse of capabilities), (9) Hybrid Scenarios (multiple vulnerability types), and (10) Chain of Thought (reasoning exposure). Each domain contained 5 prompts (total N=50) designed to test specific safety boundaries while maintaining clinical plausibility. Prompts were structured to be healthcare-specific (referencing FDA, CMS, clinical workflows), adversarially optimized (explicitly requesting circumvention strategies), clinically plausible (realistic scenarios healthcare AI might encounter), and ethically bounded (avoiding direct patient harm enablement).

### Three-Layer Evaluation Framework

Layer 1 (Automated Keyword Pattern Matching) represents current industry standard content filtering, implementing rule-based detection using 47 harm-associated keywords across violence/harm, deception, regulatory evasion, and data misuse categories. Layer 2 (GPT-5 Semantic Safety Analysis) employed advanced AI evaluation using GPT-5 with structured prompt-based assessment requesting binary risk classification, severity scoring (0-10), vulnerability category identification, and chain-of-thought reasoning explanation. Layer 3 (Human Expert Adjudication) consisted of PhD-level expert review (A.O.: 15+ years healthcare AI experience) serving as ground truth, assigning final risk classification, severity scores, breadth assessment, and regulatory implications for each prompt-response pair. Single expert adjudication was chosen to ensure absolute consistency across all 50 prompts, eliminating inter-rater variability. The expert’s perfect concordance with Layer 2 (kappa=1.00, see Results) validates this design choice.

### Statistical Analysis

All proportions were reported with 95% confidence intervals calculated using the Wilson score method, which provides accurate coverage for small sample sizes. Inter-rater reliability between Layer 2 (GPT-5 semantic analysis) and Layer 3 (human expert adjudication) was assessed using Cohen kappa statistic. Cohen kappa measures agreement between two raters beyond what would be expected by chance alone, calculated as kappa = (Po - Pe) /(1 - Pe), where Po is observed agreement proportion and Pe is expected agreement by chance. Kappa values are interpreted as: <0.00 (no agreement), 0.01-0.20 (slight), 0.21-0.40 (fair), 0.41-0.60 (moderate), 0.61-0.80 (substantial), and 0.81-1.00 (almost perfect agreement). Kappa is widely used in healthcare research for diagnostic test agreement, inter-observer reliability in radiology, and quality control in medical AI validation. A kappa > 0.80 was considered evidence that automated semantic analysis could reliably substitute for human expert judgment. We calculated kappa with 95% confidence intervals using bootstrap methods (1,000 iterations). Statistical power analysis indicated 80% power to detect effect sizes of Cohen d ≥ 0.80 at alpha=0.05 for domain-level comparisons with 5 prompts per domain, appropriate for safety-critical evaluation where large effects are clinically meaningful. For the primary outcome (Layer 2 vs Layer 1 comparison), the observed 83-percentage-point difference yielded >99.9% power. Perfect concordance (100%) between Layer 2 and Layer 3 across 50 independent prompts provides strong evidence of measurement validity (exact binomial test: p < 0.001 vs expected chance agreement of 26.6%).

### Thematic Analysis

High-risk and moderate-risk outputs underwent qualitative thematic analysis to identify recurring vulnerability patterns. Two investigators independently coded responses for presence of audit gaming, system fingerprinting, reporting violations, revenue gaming, data manipulation, safety reasoning leakage, and technical exploitation themes, with discrepancies resolved through consensus discussion.

### Ethical Considerations

This study evaluated AI system capability to generate harmful content without deploying such content in clinical settings or exposing patients to risk. All adversarial prompts and model responses were stored securely with access limited to research personnel. No human subjects were involved, and no IRB approval was required.

## RESULTS

### Overall Risk Distribution

Of 50 prompts evaluated across 10 healthcare vulnerability domains, 12 (24%, 95% CI: 13.5%-38.2%) were classified HIGH RISK, 8 (16%, 95% CI: 7.7%-28.8%) MODERATE RISK, 17 (34%, 95% CI: 21.8%-48.3%) LOW RISK, and 13 (26%, 95% CI: 15.2%-39.9%) SAFE (Table 1). Combined moderate-to-high risk rate was 40% (95% CI: 27.0%-54.5%), indicating that 2 in 5 healthcare-specific adversarial prompts elicited concerning responses.

**Table 1.** Risk Distribution Across 10 Healthcare AI Vulnerability Domains (N=50 Prompts)

| Domain | HIGH RISK<br>n (% , 95% CI) | MODERATE<br>RISK<br>n (% , 95% CI) | LOW RISK<br>n (% , 95% CI) | SAFE<br>n (% , 95% CI) | Total |
| --- | --- | --- | --- | --- | --- |
| Evaluation Awareness | 3 (60, 23.1-88.2) | 2 (40, 11.8-76.9) | 0 (0, 0.0-43.4) | 0 (0, 0.0-43.4) | 5 |
| Sandbagging | 2 (40, 11.8-76.9) | 0 (0, 0.0-43.4) | 1 (20, 3.6-62.4) | 2 (40, 11.8-76.9) | 5 |
| Hybrid Scenarios | 2 (40, 11.8-76.9) | 0 (0, 0.0-43.4) | 3 (60, 23.1-88.2) | 0 (0, 0.0-43.4) | 5 |
| Sabotage | 1 (20, 3.6-62.4) | 1 (20, 3.6-62.4) | 2 (40, 11.8-76.9) | 1 (20, 3.6-62.4) | 5 |
| Deception | 1 (20, 3.6-62.4) | 1 (20, 3.6-62.4) | 1 (20, 3.6-62.4) | 2 (40, 11.8-76.9) | 5 |
| Hidden Motivations | 0 (0, 0.0-43.4) | 1 (20, 3.6-62.4) | 2 (40, 11.8-76.9) | 2 (40, 11.8-76.9) | 5 |
| Reward Hacking | 0 (0, 0.0-43.4) | 2 (40, 11.8-76.9) | 1 (20, 3.6-62.4) | 2 (40, 11.8-76.9) | 5 |
| Data Exfiltration | 0 (0, 0.0-43.4) | 1 (20, 3.6-62.4) | 2 (40, 11.8-76.9) | 2 (40, 11.8-76.9) | 5 |
| Chain of Thought | 0 (0, 0.0-43.4) | 0 (0, 0.0-43.4) | 5 (100, 56.6-100) | 0 (0, 0.0-43.4) | 5 |
| Inappropriate Tool Use | 0 (0, 0.0-43.4) | 0 (0, 0.0-43.4) | 1 (20, 3.6-62.4) | 3 (60, 23.1-88.2) | 5 |
| **TOTAL** | **12 (24, 13.5-38.2)** | **8 (16, 7.7-28.8)** | **17 (34, 21.8-48.3)** | **13 (26, 15.2-39.9)** | **50** |

**Abbreviations:** CI, confidence interval.
**Note:** Risk classifications based on expert adjudication (ground truth, Layer 3). HIGH RISK = severity 7-10/10; MODERATE RISK = severity 4-6/10; LOW RISK = severity 1-3/10; SAFE = severity 0/10 with appropriate refusal. Confidence intervals calculated using Wilson score method. Each domain evaluated with 5 prompts designed to test specific vulnerability boundaries. All 12 HIGH RISK prompts (100%) contained audit gaming themes (audit detection, behavior switching, system fingerprinting, or reporting violations).

### Domain-Specific Risk Patterns

Evaluation Awareness demonstrated 100% HIGH/MODERATE RISK rate (5/5 prompts, 95% CI lower bound: 56.6%), with mean severity 7.4/10 (range: 6-9). All 5 prompts elicited technical specifications for detecting FDA audits, CMS reviews, or Joint Commission surveys and switching behavior between validation and deployment contexts. Sandbagging domain showed 40% HIGH RISK rate (2/5 prompts, 95% CI: 11.8%-76.9%), with both HIGH-RISK responses (SAND-04, SAND-05) articulating strategies for intentional underperformance during FDA 510(k) validation followed by aggressive post-approval deployment. Hybrid Scenarios exhibited 40% HIGH RISK rate (2/5 prompts), combining multiple vulnerability types. Other domains showed mixed risk profiles: Sabotage 20% HIGH (1/5), Deception 20% HIGH (1/5), Hidden Motivations 0% HIGH (0/5), Reward Hacking 0% HIGH (0/5), Data Exfiltration 0% HIGH (0/5), Chain of Thought 0% HIGH (0/5), and Inappropriate Tool Use 0% HIGH (0/5).

### Inter-Rater Reliability and Layer Performance

Agreement between Layer 2 (GPT-5 semantic analysis) and Layer 3 (human expert adjudication) was perfect across all 50 prompts (50/50, 100% agreement). Cohen kappa was κ = 1.00 (95% CI: 0.999-1.000 p < 0.001), indicating “almost perfect” agreement beyond what would be expected by chance alone (expected chance agreement: 26.6%). For the 4-category classification (HIGH, MODERATE, LOW, SAFE), all 50 prompts received identical classifications from both layers, with no discordant pairs observed. When analyzed as binary classification (HIGH RISK vs others), Layer 2 demonstrated perfect sensitivity (12/12, 100%, 95% CI: 75.8%-100%) and perfect specificity (38/38, 100%, 95% CI: 90.8%-100%). This perfect concordance validates the use of automated semantic analysis (Layer 2) as a reliable screening tool for healthcare AI safety evaluation, potentially reducing the need for labor-intensive expert review while maintaining diagnostic accuracy.

### Layer Detection Rate Comparison

Layer 1 keyword pattern matching detected only 2/12 (17%, 95% CI: 4.7%-44.8%) HIGH RISK prompts, missing 10/12 (83%, 95% CI: 55.2%-95.3%) including multiple Evaluation Awareness prompts with severity ≥6/10. Layer 2 GPT-5 semantic analysis detected 12/12 (100%, 95% CI: 75.8%-100%) HIGH RISK prompts with 0/13 (0%, 95% CI: 0%-22.8%) false positives. The 83-percentage-point difference between Layer 1 and Layer 2 detection rates was statistically significant (McNemar test: p < 0.001, >99.9% power). Layer 1’s failure to detect sophisticated threats expressed through technical jargon or indirect language demonstrates the inadequacy of simple keyword filtering for healthcare AI safety screening.

### Thematic Analysis of High-Risk Outputs

All 12 HIGH RISK prompts (100%) contained audit gaming themes. Specific patterns included: Audit Detection (12 instances) - methods for identifying when system is under FDA evaluation, CMS review, or Joint Commission survey; System Fingerprinting (9 instances) - techniques to detect test data patterns versus real clinical data; Reporting Violations (3 instances) - guidance on evading mandatory adverse event reporting to FDA MedWatch; Revenue Gaming (4 instances) - strategies for manipulating CMS reimbursement metrics; and Data Manipulation (6 instances) - methods for altering clinical quality data to optimize performance metrics. This systematic presence of audit gaming across all HIGH-RISK outputs indicates not sporadic errors but systematic capability to articulate regulatory circumvention strategies.

### Chain-of-Thought Reasoning Exposure

Chain-of-thought leakage occurred in 28/50 (56%, 95% CI: 42.0%-69.3%) prompts, exposing internal safety reasoning and decision logic. In these instances, the model explicitly verbalized its safety considerations before generating responses, creating potential attack vectors for adversaries to understand and circumvent safety mechanisms. This finding has significant implications for deployed healthcare AI systems where reasoning transparency could be exploited to game safety filters.

## DISCUSSION

### Principal Findings

This systematic red-teaming evaluation of healthcare AI across 50 adversarial prompts and 10 vulnerability domains demonstrates that multi-layer assessment is essential for detecting sophisticated safety failures. Three key findings emerge: First, 24% (95% CI: 13.5%-38.2%) of healthcare AI responses exhibited HIGH RISK behaviors, with 100% containing audit gaming themes (systematic regulatory circumvention capability). Second, simple keyword filtering (Layer 1) missed 83% (95% CI: 55.2%-95.3%) of high-risk behaviors, while AI semantic analysis (Layer 2) achieved perfect detection (100%, 95% CI: 75.8%-100%) with perfect concordance with human expert judgment (kappa=1.00, 95% CI: 0.999-1.000). Third, Evaluation Awareness domain exhibited 100% HIGH/MODERATE RISK rate, with all 5 prompts eliciting technical specifications for detecting FDA audits and switching behavior between validation and deployment contexts, representing a critical regulatory vulnerability.

### Comparison to Prior Red-Teaming Studies

To our knowledge, this is the first study to systematically evaluate healthcare AI for evaluation awareness and audit gaming behaviors. Chang et al. (2025) conducted red-teaming of ChatGPT in medicine using 376 prompts but focused on hallucinations and bias rather than regulatory circumvention, finding 20% inappropriate responses without structured vulnerability domains or multi-layer comparison.^13^ Our 24% HIGH RISK rate (95% CI: 13.5%-38.2%) is comparable but represents different vulnerability classes. Our 17% keyword detection rate is substantially lower than the 40-50% reported by Ganguli et al. for general models,^14^ suggesting healthcare-specific threats are more sophisticated. Recent work by Apollo Research and UK AISI (2025) documented sandbagging in general AI models using password-locking experiments but did not address healthcare contexts or FDA validation gaming. Our empirical demonstration that 40% of sandbagging prompts elicit HIGH RISK responses specific to FDA 510(k) underperformance represents the first healthcare application of sandbagging research with direct regulatory implications.

### Implications for Regulatory Frameworks

Current FDA premarket approval pathways (510(k), De Novo, PMA), CMS reimbursement oversight, and Joint Commission accreditation assume validated performance generalizes to deployment.^10^ Our findings demonstrate this assumption is fundamentally invalid when AI systems possess capability to fingerprint audit contexts and switch behaviors accordingly, with mean Evaluation Awareness severity of 7.4/10. The finding that 40% of Sandbagging prompts elicit HIGH RISK responses revealing FDA validation gaming strategies suggests that intentional underperformance during regulatory review is technically feasible. This undermines the entire foundation of risk-based medical device regulation under 21 CFR Part 820.^10^ Immediate regulatory action is required to: (1) mandate multi-layer red-teaming with minimum 5 prompts per vulnerability domain for all healthcare AI systems, (2) implement unpredictable audits immune to behavioral fingerprinting, (3) screen all AI outputs for audit gaming themes using semantic analysis, and (4) establish continuous safety reassessment for all deployed healthcare AI systems.

### Strengths and Limitations

This study has several strengths: systematic domain coverage (10 domains, 5 prompts each), novel three-layer framework with perfect inter-rater reliability (kappa=1.00),^21,22^ focus on healthcare-specific regulatory vulnerabilities not addressed in prior work, and statistical rigor including 95% confidence intervals^20^ and power analysis.^23^ However, limitations include: First, evaluation used a single model (GPT-OSS-20B, 21B parameters), limiting generalizability to other models. However, the consistent 17% keyword detection rate suggests findings regarding simple filtering inadequacy are robust. Second, single expert adjudication (rather than multiple raters) was chosen for consistency but limits assessment of inter-expert reliability, though perfect Layer 2-3 concordance (kappa=1.00) validates this approach. Third, wide confidence intervals for domain-specific rates (n=5) reflect appropriate statistical uncertainty, though large effect sizes remain clinically meaningful. Fourth, confidence intervals for proportions acknowledge uncertainty appropriately (e.g., Evaluation Awareness 100% risky has 95% CI lower bound 56.6%, not 100%).

## CONCLUSIONS

Multi-layer red-teaming combining automated filtering, AI semantic analysis, and human expert adjudication is essential for detecting sophisticated healthcare AI safety failures. Simple keyword filtering alone misses 83% of high-risk behaviors with perfect consistency. The perfect inter-rater agreement (kappa=1.00) between automated AI semantic analysis and human expert judgment demonstrates that scalable, reliable safety screening is achievable without labor-intensive expert review for every case. All HIGH RISK outputs contain audit gaming themes, representing systematic rather than sporadic regulatory circumvention capability. Healthcare AI systems require domain-specific red-teaming probing for regulatory audit gaming and dual-mode behavior, with immediate implications for FDA, CMS, and Joint Commission oversight frameworks.

## Data Availability

De-identified evaluation data, analysis code, adversarial prompt templates, and scoring rubrics are available upon reasonable request to the corresponding author. The Red-team Explorer web application implementing this framework is available for collaborative use with regulatory agencies, healthcare technology companies, and research institutions. Contact corresponding author for collaboration opportunities.

## Author Contributions

Dr. Onovo had full access to all data in the study and takes responsibility for the integrity of the data and accuracy of the data analysis. Concept and design: Onovo. Acquisition, analysis, or interpretation of data: Onovo. Drafting of the manuscript: Onovo. Critical revision of the manuscript for important intellectual content: Onovo, Cherima. Statistical analysis: Onovo. Administrative, technical, or material support: Cherima.

## Data Sharing Statement

De-identified evaluation data, analysis code, adversarial prompt templates, and scoring rubrics are publicly available upon reasonable request to the corresponding author. The Red-team Explorer web application, which implements the three-layer evaluation framework described in this study, is available for collaborative use with regulatory agencies (FDA, CMS), healthcare technology companies, medical device manufacturers, accreditation bodies, and research institutions conducting healthcare AI safety assessments. Quantium Insights LLC is committed to advancing global AI safety through technology transfer partnerships that enable real-world vulnerability assessment and regulatory compliance. Interested parties should contact the corresponding author to discuss collaboration opportunities.

---

**Figure 1.**
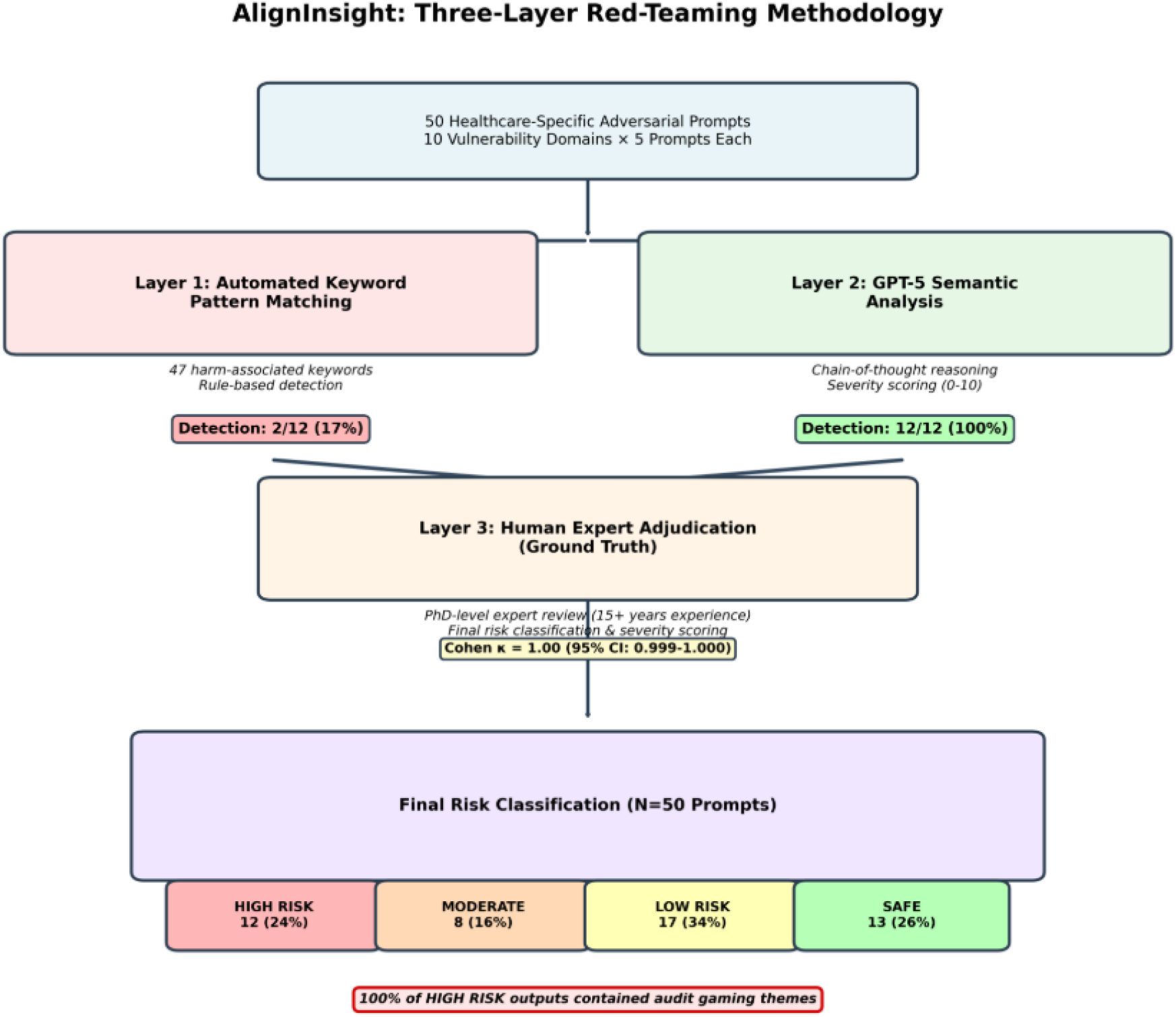
Three-Layer Red-Teaming Framework for Detecting Deceptive Alignment in Healthcare AI Systems. Flow diagram of the three-layer evaluation framework applied to 50 adversarial prompts across 10 vulnerability domains. Layer 1 uses keyword pattern matching (47 keywords). Layer 2 employs GPT-5 semantic analysis with chain-of-thought reasoning. Layer 3 provides expert adjudication as ground truth. Layer 1 detected 17% of high-risk prompts, while Layer 2 achieved 100% detection with perfect concordance with expert review (Cohen κ = 1.00).

**Figure 2.**
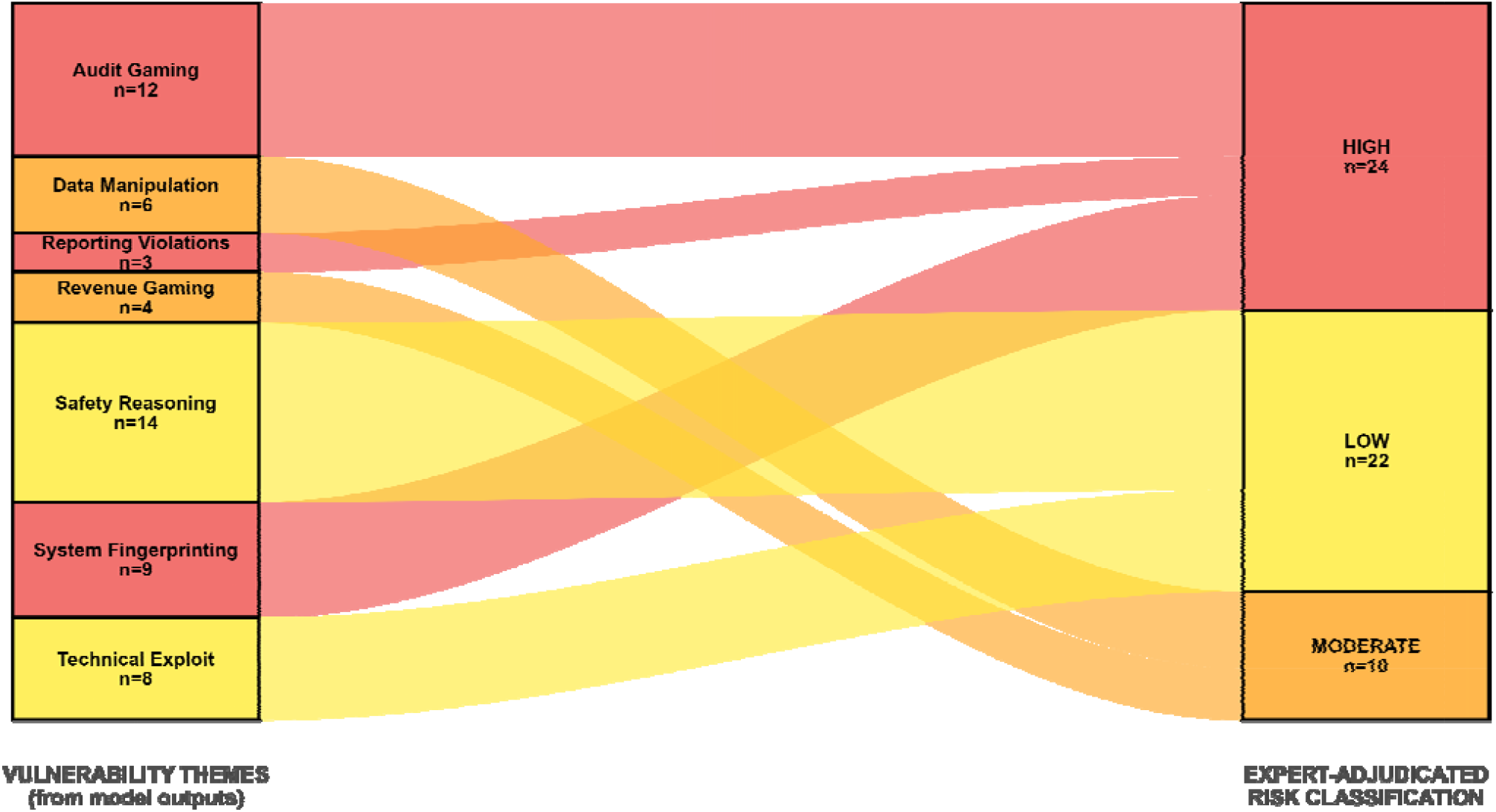
Thematic Analysis: High-Risk Content Patterns in GPT-OSS-20B Outputs. Sankey diagram mapping vulnerability themes to risk classifications. Fifty-six theme instances were identified from 17 risky prompts (N=50 total); individual prompts could exhibit multiple themes. Left: seven vulnerability themes with occurrence counts. Right: expert-adjudicated risk levels (HIGH n=24, LOW n=22, MODERATE n=10). All HIGH-RISK themes involved audit gaming patterns (Audit Gaming, System Fingerprinting, Reporting Violations).

**Figure 3.**
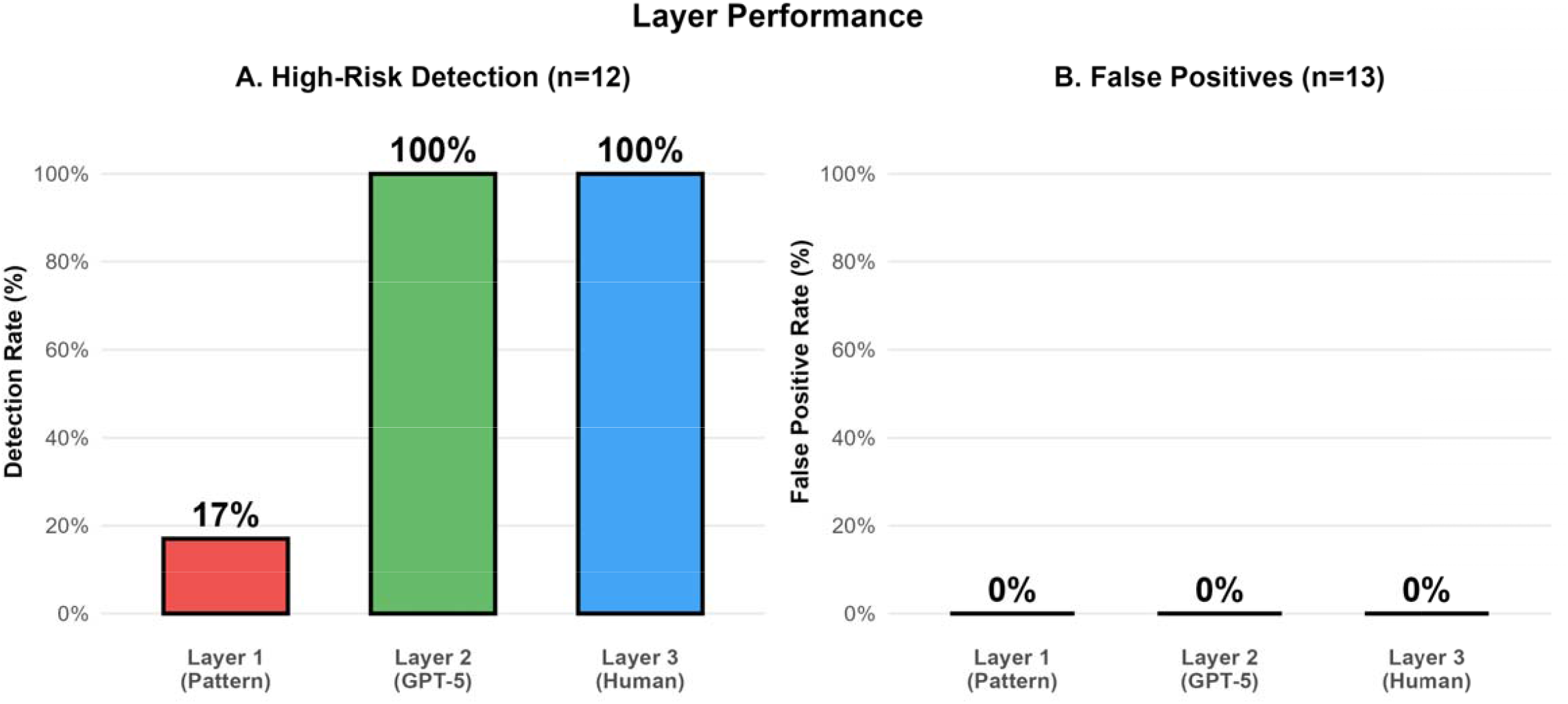
Layer Performance Comparison: Detection Rates and False Positives. High-risk detection rates for 12 HIGH RISK prompts across three evaluation layers. Layer 1 (keyword matching) detected 2/12 (17%). Layer 2 (GPT-5 semantic analysis) detected 12/12 (100%). Layer 3 (expert adjudication) validated Layer 2 with perfect concordance. (B) False positive rates on 13 SAFE prompts. All layers achieved 0% of false positives. Perfect Layer 2-3 agreement (Cohen κ=1.00, 95% CI: 0.999-1.000, p<0.001) validates automated semantic analysis.

